# Structural Brain Networks and the Seizure-Survival Paradox in Glioblastoma

**DOI:** 10.64898/2026.09.04.26362299

**Authors:** Daniel J. Zhou, Marc Jaskir, Julian Gal, Yombe Fonkeu, Alfredo Lucas, Quy Cao, Christos Davatzikos, MacLean P. Nasrallah, Jose Garcia, Rubo Xing, Richard E. Phillips, Colin A. Ellis, Sandhitsu Das, James J. Gugger, Nishant Sinha, Joel M. Stein, Kathryn A. Davis

**Author notes:** **Corresponding Author:** Kathryn Adamiak Davis, MD, MSc, FANA, FAES University of Pennsylvania, 301 Hayden Hall, Philadelphia, PA 19104.

## Abstract

**Background:** In glioblastoma, tumor-related epilepsy (TRE) is paradoxically associated with longer survival, but the mechanism is unclear. We hypothesized that structural connectivity is associated with seizure susceptibility and survival, and tested whether these associations are independent or mediated.

**Methods:** We retrospectively studied glioblastoma patients with preoperative diffusion-weighted MRI. Fractional anisotropy-weighted connectivity matrices were constructed using lesion-aware tractography. Mean edge weight and local efficiency were computed per Yeo 7-network and with global efficiency at hemisphere and whole-brain levels. Associations with TRE, overall survival (OS), and progression-free survival (PFS) were assessed using logistic regression and Cox proportional hazards models with false discovery rate correction. Causal mediation analysis tested whether connectivity and TRE mediate each other’s relationship with overall survival.

**Results:** We included 420 glioblastoma patients. Of 406 classifiable for TRE, 217 (53%) had TRE. TRE was associated with higher local efficiency across all seven contralateral networks (*d=*0.42-0.49, *q<*0.010) and five of seven ipsilateral networks (*d*=0.44-0.49, *q<*0.050). In adjusted models, higher contralateral global efficiency was associated with longer OS (HR 0.84, *q=*0.021), with no metric associated with PFS. TRE did not mediate the connectivity-OS relationship. Contralateral local efficiency partially mediated the TRE-OS association when postoperative KPS was excluded from the model (ACME 16.6 days, 7.8% mediated, *p*=0.029), though this was attenuated when KPS was included (*p*=0.074).

**Conclusions:** Structural connectivity may partially mediate the seizure-survival association, suggesting preserved network architecture may contribute to longer survival in glioblastoma patients with TRE, though most of the effect operates through other pathways.

## Introduction

Seizures are among the most common neurological manifestations of glioblastoma, occurring in approximately 25-35% of patients at initial diagnosis and with a cumulative incidence of over half throughout the disease course.^1–3^ Particularly in patients with brain tumors, seizures significantly impair quality of life across physical, cognitive, and psychosocial domains.^1,4^ Multiple studies have shown that patients with glioblastoma who develop seizures experience longer overall survival (OS), even after adjustments for age, performance status, extent of resection, tumor location and volume, and *MGMT* promoter methylation status, and experience longer progression-free survival (PFS), though evidence for PFS has been more limited.^2,5–8^ The mechanisms underlying this seizure-survival association remain unclear. Proposed explanations include lead-time bias from earlier diagnosis or distinct biological features of epileptogenic tumors, though the structural neurobiological basis for this relationship has yet to be investigated.^2,5,6^

Structural brain network analysis using diffusion-weighted imaging (DWI) offers a framework for characterizing how gliomas disrupt white matter networks. Prior work has demonstrated that glioblastoma causes widespread connectome disruptions as inferred by decreases of fractional anisotropy (FA). These disruptions are associated with impaired cognitive function and worse OS in glioblastoma patients. Moreover, reduced between-network structural connectivity across canonical functional brain networks maps to cognitive deficits in treated glioma patients.^9–11^ Separately, evidence from non-tumor epilepsy suggests that epileptic networks are characterized by paradoxical increases in regional structural connectivity, such as within limbic structures for temporal lobe epilepsy, despite widespread fiber loss, a pattern referred to as network regularization.^12–14^ This raises the question of whether seizure susceptibility in glioblastoma reflects a similar circuit-specific reorganization, confined to particular networks, or instead a more distributed preservation of connectivity across many networks. How this network topology relates to both seizure susceptibility and survival has not been adequately examined.

In this study, we characterized structural network alterations associated with both tumor-related epilepsy (TRE) and survival in a large cohort of glioblastoma patients using FA-weighted graph theory metrics organized by the Yeo canonical functional networks.^15^ We hypothesized that preserved structural connectivity across distributed functional networks would be associated with seizure susceptibility. We further hypothesized that greater connectivity disruption would be associated with worse OS and PFS, and explored whether connectivity and seizure susceptibility mediate each other’s relationship with survival or reflect independent prognostic pathways (Fig. 1A).

**Figure 1.**
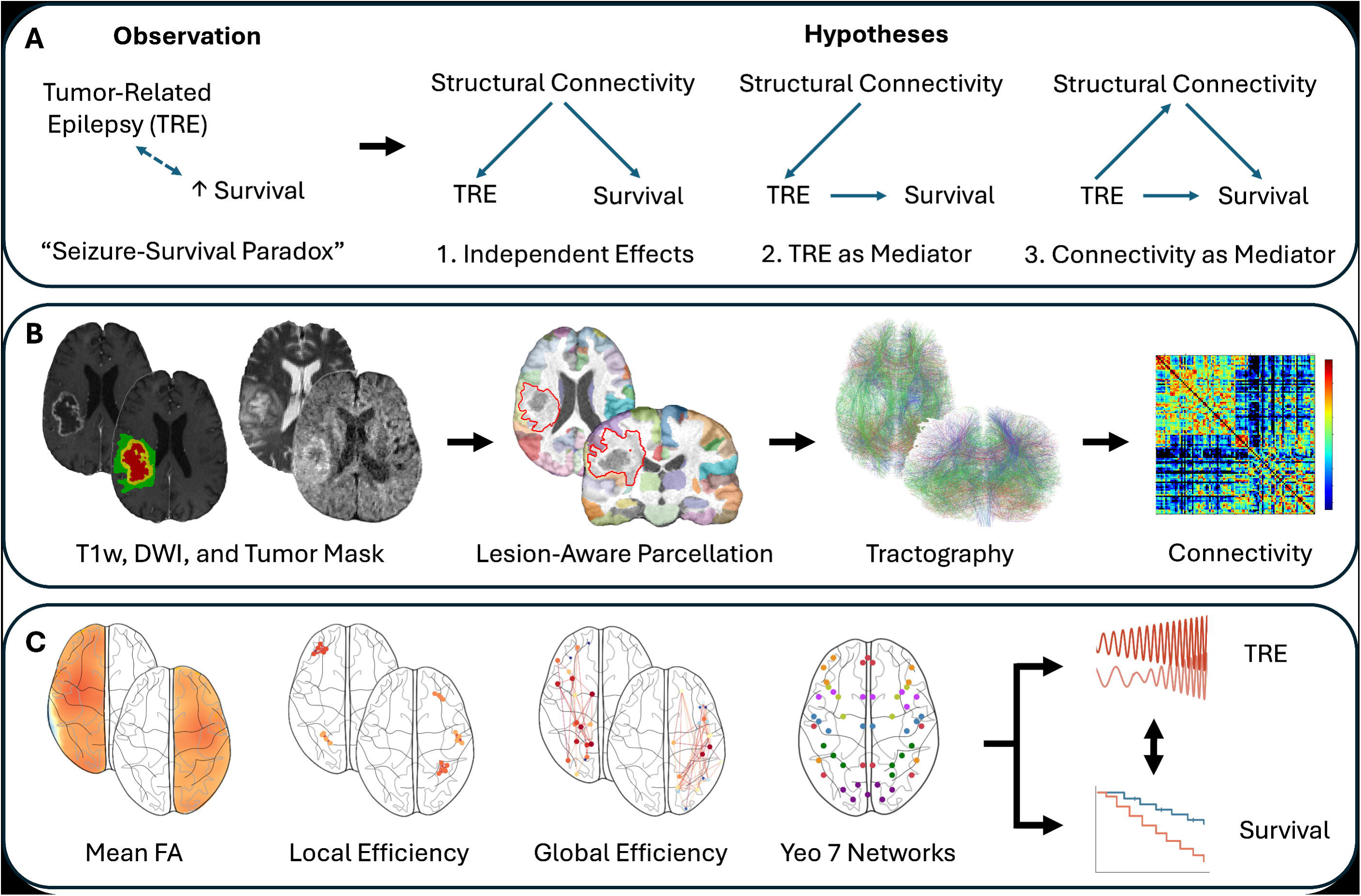
Study overview and analysis pipeline. (A) Overview of the seizure-survival paradox and hypotheses tested. (B) Structural and diffusion-weighted images were preprocessed, tumor regions were segmented, and a lesion-aware parcellation and tractography pipeline was used to generate fractional anisotropy (FA)-weighted connectivity matrices. (C) Mean FA edge weight and local efficiency were computed per Yeo network and, along with global efficiency, at the hemisphere and whole-brain level. These metrics were tested for associations with tumor-related epilepsy and survival, and counterfactual causal mediation analysis tested whether tumor-related epilepsy and connectivity mediate each other’s relationship with survival.

## Materials and Methods

### Patient Selection

This study was approved by the University of Pennsylvania Institutional Review Board (835008, 706564), and informed consent was obtained from all participants or their legally authorized representatives. Patients were included if they had a diagnosis of glioblastoma and underwent brain MRI between January 2010 and April 2023 at the University of Pennsylvania. Patient selection, molecular confirmation of glioblastoma diagnosis per 2021 WHO classification criteria, and exclusion criteria have been previously described.^8^ Briefly, all cases underwent repeat molecular testing on archived tumor tissue, with age-stratified molecular requirements for glioblastoma classification. Patients under 30 years of age, those with H3 K27 or G34 alterations or pleomorphic xanthoastrocytoma, and those who received only biopsy without resection were excluded.^16,17^ DWI scans were additionally required to have adequate metadata for preprocessing, and scans with poor DWI quality (raw neighbor correlation <0.8) were excluded.

### Data Extraction

Demographic, clinical, and imaging data were extracted by clinical neurologists (D.Z., J.G) and neurology resident physician (Y.F.) through systematic chart review. Follow-up data were current through July 21, 2025. Age was defined at the time of the diagnostic MRI. *MGMT* promoter methylation status was obtained from clinical notes or pathology reports; patients without documented testing were excluded from methylation-dependent analyses. Extent of resection was categorized as gross total, near gross total (≥95% removal), or subtotal (<95% removal) based on operative reports or, when unavailable, postoperative imaging.

Tumor-related epilepsy (TRE) was defined as new-onset seizures attributable to the brain tumor, occurring either preoperatively or more than seven days postoperatively, in the absence of identifiable acute precipitants. Patients who experienced seizures within seven days of resection, or who developed seizures in the setting of a significant postoperative complication (large intracranial hemorrhage, infection, or anoxic injury), were excluded from TRE classification. Overall survival (OS) was measured from the date of initial tumor resection to death or last follow-up. Progression-free survival (PFS) was measured from initial resection to first documented radiographic progression based on clinical radiology reports and neuro-oncology notes. Only unequivocal progression was counted as an event, while equivocal findings prompting surveillance imaging were not.

### Image Processing

All MRI scans were co-registered to the SRI24 atlas at 1mm^3^ isotropic resolution, skull-stripped, and preprocessed following the BraTS protocol as previously described.^18^ Brain tumor regions were segmented using validated deep learning algorithms, and a composite tumor mask encompassing all tumor components (enhancing tumor, necrotic core, and edema) was generated for use in lesion-aware parcellation and tractography (Fig. 1B).^18^

DWI data were acquired on Siemens scanners using a single-shell protocol (b=1000 s/mm2, 30 diffusion directions, slice thickness 3 mm), with minor acquisition variations across the cohort as previously described.^18^ DWI data were preprocessed using QSIPrep, which performed denoising, Gibbs unringing, susceptibility distortion correction, and lesion masking.^19^ Raw neighbor correlation was extracted as a measure of DWI data quality.^20^ Whole-brain tractography was performed using QSIRecon with fiber orientation distribution estimation via single-shell 3-tissue constrained spherical deconvolution (SS3T-CSD), followed by probabilistic tractography using the iFOD2 algorithm.^19^

### Structural Connectome Construction

Since FreeSurfer cortical reconstructions often fail in glioblastoma due to tumor- and edema-related distortion, cortical parcellation instead used the Schaefer 100-parcel atlas, organized according to the Yeo 7-network parcellation (visual, somatomotor, dorsal attention, salience/ventral attention, limbic, frontoparietal control, and default mode).^15,21^ The atlas, defined in MNI152NLin2009cAsym space, was warped into each subject’s native (ACPC) space by applying the inverse of the QSIPrep-derived MNI-to-T1w transform using ANTs.^19,22,23^ Subcortical regions (15 ROIs) were segmented using SynthSeg, a deep learning tool robust to pathological anatomy.^24^ The brainstem ROI was excluded from analyses due to known tractography artifacts and incomplete segmentation of the brainstem in a subset of subjects.

The cortical and subcortical parcellations were combined to produce 114 total ROIs. Dominant tumor lobe was classified as frontal, temporal, parietal, or other (occipital, insula, cingulate, or subcortical), based on the predominant regional volume of enhancing tumor as previously described.^8^ Tumors were additionally classified as predominantly cortical or subcortical based on the proportion of enhancing tumor volume overlapping subcortical parcellation ROIs, with tumors exceeding 50% subcortical overlap designated as predominantly subcortical.

The tumor segmentation, resampled from native T1w to ACPC space via affine transformation, was used to set all overlapping parcellation voxels to label 0, preventing any streamline endpoints from falling within the tumor. Streamlines passing through the tumor/edema mask were then removed from the tractogram using tckedit with an exclusion mask.^25^ FA-weighted 114×114 connectivity matrices were generated using tck2connectome with the combined atlas.^25^ Per-network analyses were restricted to the 100 cortical ROIs, and subcortical ROIs were included in hemisphere-level and whole-brain graph metrics. Tumor laterality was determined by counting tumor- and edema-mask voxels overlapping parcellation ROIs in each hemisphere; the hemisphere with greater overlap was designated tumor-dominant, and ipsilateral/contralateral hemispheres were defined accordingly.

### Quality Control

Two subjects with raw neighbor correlation below 0.80 were excluded for poor DWI quality. An additional five subjects were found to have missing right accumbens parcellations, two of whom were also missing the right amygdala. Of these, three had large tumors causing severe deformation of the affected ROIs due to direct mass effect, so their missing edges were zero-padded to reflect absent measurable connectivity, a conservative assumption that may underestimate true connectivity in regions with severe deformation. The remaining two were excluded because SynthSeg did not return the affected ROI in the contralateral hemisphere, away from the tumor, where a missing label likely reflected segmentation failure rather than true anatomical loss and therefore could not be justifiably zero-padded.

Parcellation overlays and whole-brain tractography projections were generated as 2D images for the 50 subjects with the largest total tumor and edema volumes (range: 146–237 cm^3^). Parcellation images show axial, coronal, and sagittal slices through the tumor centroid; tractography images show planar projections of the full streamline set. Images were visually reviewed (D.Z.) to confirm that lesion-masked atlases correctly excluded tumor voxels while preserving surrounding cortex and subcortex, and that lesion-filtered tractography showed plausible streamline coverage. All 50 subjects passed review.

### Graph Theory Metrics

Graph theory provides a mathematical framework for characterizing the topology of brain networks.^26^ Local efficiency quantifies how well a node’s neighbors are interconnected, reflecting the capacity for specialized processing within a network neighborhood. Global efficiency measures the ease of information transfer across the entire network, reflecting overall network integration. These metrics were selected because they capture complementary aspects of network organization, specifically local segregation and global integration, that have been shown to be affected in both glioma and epilepsy.^9,12^

Graph theory metrics were extracted from the FA-weighted connectivity matrices for ipsilateral, contralateral, and whole-brain regions (Fig. 1C). Raw FA edge weights were used, and zero-weight entries were treated as absent edges. For each of the seven Yeo networks, local efficiency was computed as the mean nodal local efficiency across the network’s ROIs, using a geometric-mean formulation.^27^ At the hemisphere and whole-brain level, mean local efficiency and global efficiency were computed. Global efficiency was calculated as the mean inverse shortest-path length across all node pairs, with FA edge weights transformed to distances (d = 1/FA) before shortest-path computation.^28^ Mean FA edge weight was also extracted at the network and hemisphere level as a direct measure of white matter microstructural integrity.

Tumor-affected ROIs were not excluded from graph metric computations. The lesion-aware tractography pipeline removed streamlines overlapping with tumor masks to allow perilesional ROIs to contribute their connectivity information, consistent with prior approaches in glioma connectomics.^10,29^

### Statistical Analyses

Seizure and survival analyses were performed in Python 3.12 (lifelines, statsmodels, scipy). Mediation analysis was performed in R using the mediation package. Statistical significance was set at two-sided *p<*0.05 after false discovery rate (FDR) correction when applicable.

Demographic and clinical associations with TRE were assessed using Mann-Whitney U tests for continuous variables and chi-squared tests for categorical variables. Associations with OS and PFS were assessed using log-rank tests for categorical variables and univariate Cox proportional hazards regression for continuous variables. Kaplan-Meier estimates were used to report median survival with interquartile ranges.

Associations between connectivity metrics and TRE were assessed using logistic regression, and unadjusted effect sizes for TRE associations were quantified using Cohen’s *d*. Cox proportional hazards regression was used to assess the association between connectivity metrics and OS and PFS. Models for TRE analyses were adjusted for age, sex, tumor/edema volume, enhancing tumor location, and DWI quality. Models for survival were additionally adjusted for extent of resection, *MGMT* promoter methylation status, and postoperative Karnofsky Performance Status (KPS, ≥70 vs. <70). FDR correction was applied using the Benjamini-Hochberg procedure within three analysis families: per-network (mean FA and local efficiency combined per hemisphere), hemisphere-level (all three metrics across both hemispheres), and whole-brain (all three metrics). The proportional hazards assumption was verified using Schoenfeld residual tests; no violations were observed for FDR-corrected significant results.

Counterfactual causal mediation analysis was performed in two directions. The forward model tested whether TRE mediates the association between contralateral connectivity and OS, with connectivity as the exposure and TRE as a binary mediator. The reverse model tested whether contralateral connectivity mediates the association between TRE and OS, with TRE as the exposure and connectivity as a continuous mediator. The mediator model was logistic regression (forward) or linear regression (reverse), adjusted for TRE covariates, and the outcome model was a Weibull accelerated failure time (AFT) model adjusted for survival covariates.

Bootstrap inference with 2000 iterations was used to estimate the average causal mediation effect (ACME) and proportion mediated. Because postoperative KPS may reflect the functional consequences of network integrity rather than act solely as an independent confounder, adjusting for it could bias the mediation estimate toward the null. This concern is supported by the established association between connectome disruption and worse KPS in glioblastoma and the correlation between KPS and contralateral connectivity in our cohort (Supplementary Fig. 1). We therefore prespecified two outcome-model specifications reflecting alternative causal assumptions about postoperative KPS. One adjusted for KPS as a confounder, while the other excluded it as a potential intermediate variable. As postoperative KPS reflects functional status that may itself depend on connectivity, we report both specifications.

## Results

A total of 420 patients with glioblastoma were included in the study. Median age was 65 years (IQR 59-72), and 154 (37%) were female. Median OS was 428 days (IQR 211-707), and median PFS was 164 days (IQR 101-307). Fourteen patients were excluded from TRE classification but retained in survival analyses: seizures within 7 days of resection (*n*=5), or seizures in the setting of postoperative complications including large intracranial hemorrhage (*n*=6), brain abscess or ventriculitis (*n*=2), and anoxic brain injury (*n*=1). Of the remaining 406 patients classifiable for TRE, 217 (53%) had TRE, with 127 (31%) presenting with seizures at diagnosis. In those with TRE, 105 (48%) had focal-to-bilateral tonic-clonic seizures and 28 (13%) had drug-resistant epilepsy, defined by the International League Against Epilepsy as failure of adequate trials of two tolerated, appropriately chosen antiseizure medications to achieve sustained seizure freedom.^30^

Demographic and clinical characteristics in association with TRE and survival are provided in Table 1. The presence of TRE was associated with younger age (*p<*0.001) and higher postoperative KPS (*p=*0.049). Lobar tumor location was not associated with TRE (*p*=0.521), and the tumors were predominantly cortical, with only 3.3% predominantly subcortical by volume. Longer OS was associated with younger age, *MGMT* methylation, greater extent of resection, and greater postoperative KPS (all *p<*0.001). Longer PFS was associated with female sex (*p=*0.007), *MGMT* methylation (*p<*0.001), and greater extent of resection (*p=*0.005).

**Table 1:** Demographic and clinical characteristics by tumor-related epilepsy and survival.

| Categorical Variable | Tumor-Related Epilepsy |  |  | Overall Survival |  | Progression-Free Survival |  |
| --- | --- | --- | --- | --- | --- | --- | --- |
|  | Present, n (%) | No Seizure, n (%) | p-value | Median (IQR) | p-value | Median (IQR) | p-value |
| Sex |  |  | 0.555 |  | 0.585 |  | **0.007** |
| Female | 83 (38%) | 66 (35%) |  | 425 (173-681) |  | 182 (102-412) |  |
| Male | 134 (62%) | 123 (65%) |  | 428 (222-721) |  | 157 (101-270) |  |
| Tumor location |  |  | 0.521 |  | 0.059 |  | 0.270 |
| Frontal | 43 (20%) | 35 (19%) |  | 431 (213-882) |  | 162 (96-412) |  |
| Temporal | 74 (34%) | 60 (32%) |  | 524 (302-749) |  | 186 (105-339) |  |
| Parietal | 53 (24%) | 41 (22%) |  | 428 (224-648) |  | 158 (93-301) |  |
| Other | 47 (22%) | 53 (28%) |  | 316 (125-582) |  | 132 (98-232) |  |
| MGMT methylation |  |  | 0.272 |  | **<0.001** |  | **<0.001** |
| Methylated | 85 (45%) | 63 (38%) |  | 578 (263-1125) |  | 170 (98-564) |  |
| Unmethylated | 106 (55%) | 102 (62%) |  | 367 (205-564) |  | 157 (99-254) |  |
| Extent of resection |  |  | 0.734 |  | **<0.001** |  | **0.005** |
| Gross total | 88 (41%) | 72 (38%) |  | 552 (329-868) |  | 196 (112-386) |  |
| Near gross total | 47 (22%) | 47 (25%) |  | 423 (222-620) |  | 168 (93-329) |  |
| Subtotal | 82 (38%) | 70 (37%) |  | 316 (121-519) |  | 132 (55-232) |  |
| Postoperative KPS |  |  | **0.049** |  | **<0.001** |  | 0.094 |
| ≥70 | 154 (75%) | 113 (65%) |  | 524 (334-824) |  | 171 (104-315) |  |
| <70 | 51 (25%) | 60 (35%) |  | 223 (98-449) |  | 142 (48-268) |  |

| Continuous Variable | Tumor-Related Epilepsy |  |  | Overall Survival |  | Progression-Free Survival |  |
| --- | --- | --- | --- | --- | --- | --- | --- |
|  | Median (IQR) | Median (IQR) | p-value | HR (95% CI) | p-value | HR (95% CI) | p-value |
| Age (per 10 years) | 63 (57-70) | 67 (61-75) | **<0.001** | 1.33 (1.20-1.49) | **<0.001** | 1.02 (0.92-1.14) | 0.650 |
| Tumor/edema volume (per 10 cm<sup>3</sup>) | 55 (29-99) | 87 (57-121) | **<0.001** | 1.02 (1.00-1.04) | 0.135 | 1.00 (0.98-1.02) | 0.939 |

**Abbreviations:** IQR, interquartile range. KPS, Karnofsky Performance Status.

Due to missing MGMT or KPS data, fully adjusted survival analyses included 342 patients for OS and 337 for PFS. Patients missing KPS data (*n=*29) were older and had shorter OS (median 72 vs 65 years, *p=*0.002; median 224 vs 438 days, *p=*0.002), while patients missing *MGMT* methylation status (*n=*53) showed no demographic or outcome differences. The presence of TRE was associated with longer OS (HR 0.61, 95% CI 0.48-0.78, *p<*0.001) but not PFS (HR 0.88, 95% CI 0.68-1.15, *p=*0.349), after adjusting for age, sex, tumor volume, tumor location, *MGMT* methylation status, extent of resection, postoperative KPS, and DWI quality. Graph theory measures derived from FA-weighted connectivity were negatively correlated with age and tumor volume and positively correlated with extent of resection and KPS, supporting their inclusion as covariates in adjusted models (Supplementary Fig. 1).

In adjusted models, TRE was associated with higher FA-weighted connectivity across multiple metrics and networks (Fig. 2). For mean FA, TRE patients had higher values in the contralateral dorsal attention (*d=*0.27, *q=*0.026) and frontoparietal (*d=*0.29, *q=*0.037) networks with no significant ipsilateral associations (Fig. 2A-B). At the hemisphere level, whole-brain (*d=*0.36, *q=*0.008) and ipsilateral (*d=*0.34, *q=*0.038) mean FA were higher in TRE patients, but contralateral hemisphere mean FA was not (Fig. 2E). For local efficiency, TRE patients had higher values across five of seven ipsilateral networks (*d=*0.44-0.49, *q<*0.050; Fig. 2C) and all seven contralateral networks (*d=*0.42-0.49, *q<*0.010; Fig. 2D). Mean local efficiency was elevated across ipsilateral hemisphere, contralateral hemisphere, and whole-brain regions (*d=*0.43-0.49, *q=*0.008-0.021; Fig. 2F). Whole-brain global efficiency was also higher in TRE patients (*d=*0.42, *q=*0.022), but not in isolated ipsilateral and contralateral hemispheres (Fig. 2G). Unadjusted models showed stronger and more widespread associations across all metrics (Supplementary Fig. 2).

**Figure 2.**
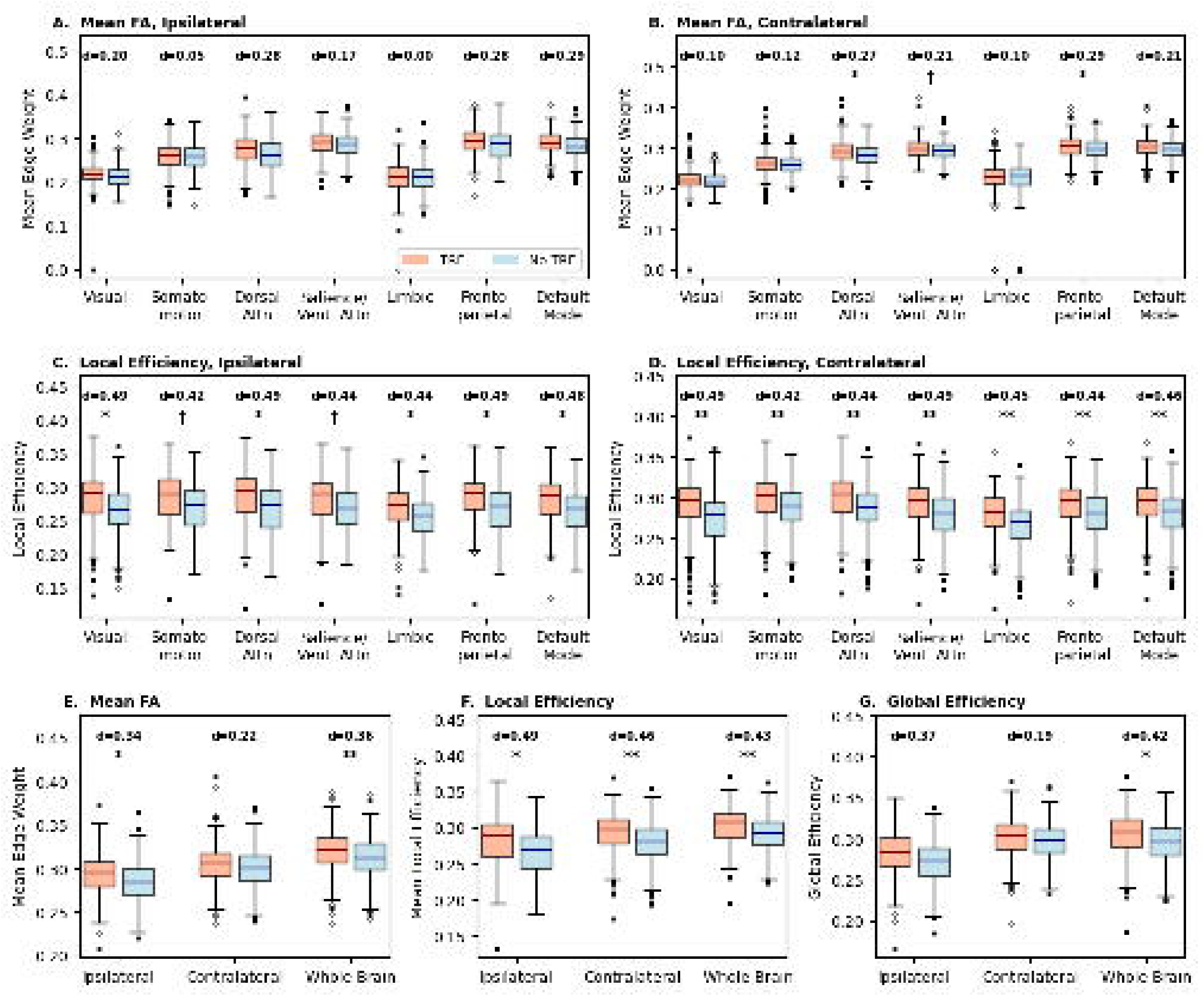
Fractional anisotropy (FA)-weighted connectivity metrics in patients with and without tumor-related epilepsy (TRE). Box plots compare distributions between patients with (red) and without TRE (blue). (A-B) Mean FA-weighted edge weight per network for ipsilateral and contralateral hemispheres. (C-D) FA-weighted local efficiency per network for ipsilateral and contralateral hemispheres. (E-G) Hemisphere-level summary metrics: (E) mean FA edge weight, (F) mean local efficiency, and (G) global efficiency for ipsilateral, contralateral, and whole-brain regions. Cohen’s d effect sizes are shown above each comparison. Significance markers from logistic regression adjusting for age, sex, tumor volume, tumor location, and DWI quality: \**q*<0.05, \*\**q*<0.01 (FDR-corrected), †*p*<0.05 (nominal).

Among patients with TRE, no connectivity metric differed between those with and without focal-to-bilateral tonic-clonic seizures, or between those with drug-resistant and drug-responsive epilepsy. Additionally, the connectivity-TRE association was consistent across preoperative and postoperative seizure subgroups, with stronger effect sizes for preoperative seizures, though neither subgroup showed significance independently (Supplementary Fig. 3).

In adjusted models, higher connectivity showed a consistent trend toward longer OS, with the strongest signals in contralateral metrics (Fig. 3). Contralateral global efficiency was significantly associated with longer OS (HR 0.84, 95% CI 0.75-0.95, *q*=0.021; Fig. 3C). No ipsilateral metric was significantly associated with OS. Unadjusted models showed substantially stronger and more widespread connectivity-OS associations (Supplementary Fig. 4). No connectivity metric was associated with PFS in either unadjusted or adjusted models (Supplementary Fig. 5).

**Figure 3.**
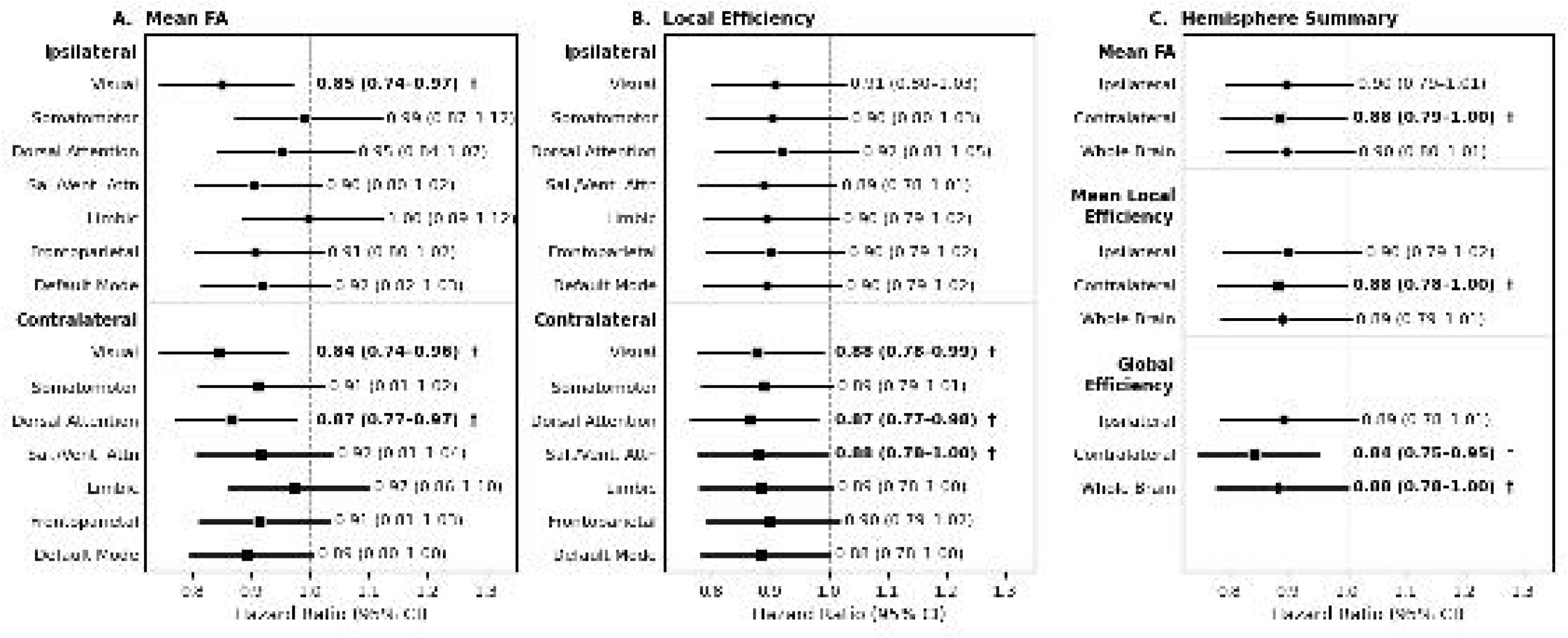
Associations between fractional anisotropy (FA)-weighted connectivity metrics and overall survival. Forest plots show hazard ratios (95% CI) from Cox proportional hazards regression adjusted for age, sex, tumor volume, tumor location, extent of resection, *MGMT* methylation status, postoperative KPS, and DWI quality. (A) Per-network mean FA edge weight. (B) Per-network local efficiency. (C) Hemisphere-level summary metrics including mean FA, mean local efficiency, and global efficiency for ipsilateral, contralateral, and whole-brain regions. Hazard ratios are per standard deviation increase in connectivity metric. \**q<*0.05 (FDR-corrected), ^†^*p<*0.05 (nominal).

Counterfactual causal mediation analysis tested whether TRE mediates the association between contralateral connectivity and OS, and whether connectivity mediates the association between TRE and OS (Fig. 4). The ACME and proportion mediated were estimated from the Weibull AFT outcome model in the complete-case sample classifiable for TRE (n=325); path hazard ratios (Fig. 4) are from Cox models refit on the same sample and covariates, are not FDR corrected, and differ slightly from the primary estimates in Fig. 3 because of the smaller sample. Testing TRE as the mediator, higher contralateral mean local efficiency was associated with TRE (OR 1.34, *p*=0.020) and TRE independently predicted longer OS (HR 0.63, *p*<0.001), but the ACME was not significant (6.3 days, 95% CI −5.7 to 34.7, *p*=0.180; proportion mediated 9.9%). The ACME remained non-significant when KPS was excluded (6.3 days, *p*=0.173; proportion mediated 7.7%; Fig. 4A-B). Testing connectivity as the mediator, TRE was associated with higher contralateral mean local efficiency (β=0.27 SD, *p*=0.019). The ACME was significant when KPS was excluded from the outcome model (16.6 days, 95% CI 1.3 to 37.9, *p*=0.029; proportion mediated 7.8%; Fig. 4D), but nonsignificant when KPS was adjusted for as a confounder (12.7 days, 95% CI −0.7 to 32.4, *p*=0.074; proportion mediated 6.1%; Fig. 4C). ACME estimates were stable across Weibull and log-normal AFT specifications.

**Figure 4.**
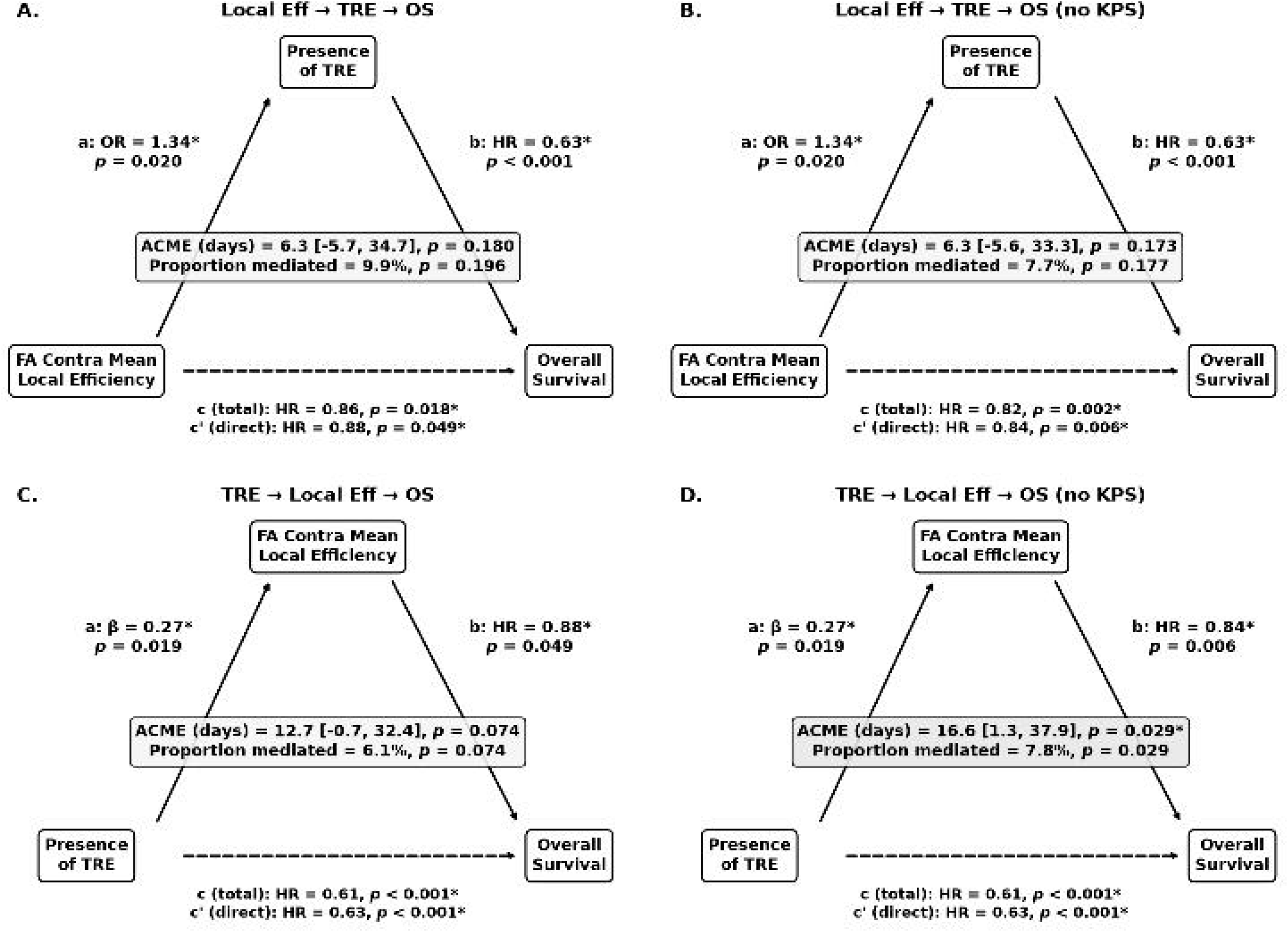
Counterfactual causal mediation analysis of the relationship between tumor-related epilepsy (TRE), fractional anisotropy (FA)-weighted contralateral mean local efficiency, and overall survival (OS). (A-B) TRE tested as the mediator of the connectivity-OS association, with (A) and without (B) postoperative KPS in the outcome model. (C-D) Contralateral mean local efficiency tested as the mediator of the TRE-OS association, with (C) and without (D) postoperative KPS. Path a: association between exposure and mediator (odds ratio for the binary mediator in A-B; standardized β for the continuous mediator in C-D). Path b: association between mediator and outcome controlling for exposure (hazard ratio). Paths c/c’: total and direct effects of the exposure on OS (hazard ratio). The average causal mediation effect (ACME) and proportion mediated are shown for each model. The outcome model was a Weibull accelerated failure time model; hazard ratios are from a Cox proportional hazards refit. Covariates were age, sex, tumor volume, extent of resection, MGMT methylation status, DWI quality, tumor location, and postoperative KPS. \**p*<0.05.

## Discussion

In this large single-center cohort of 420 patients with glioblastoma, we found that TRE was independently associated with broadly increased mean local efficiency across nearly all canonical functional networks bilaterally, while greater contralateral global efficiency was independently associated with longer OS. Mediation analysis suggested that contralateral mean local efficiency could account for a small part of the survival advantage associated with TRE. Together, these findings suggest that preserved white matter network integrity may both sustain the architecture permitting seizure generation and reflect less diffuse tumor infiltration. This positions structural connectivity as a potential correlate of the seizure-survival paradox, consistent with a model in which seizures mark rather than confer longer survival. This interpretation aligns with an emerging framework in which glioma effects are distributed across brain networks, and connectome integrity could reflect clinical outcomes.^31^ Our results extend this framework by showing that the same structural connectome measures relate to both seizure susceptibility and survival.

Peritumoral hyperexcitability driven by glutamate dysregulation, interneuron loss, and neuron-glioma synaptic interactions has been increasingly described in the glioblastoma microenvironment, yet only approximately half of patients develop clinical seizures.^32,33^ Emerging evidence suggests that seizure propagation is shaped by the underlying structural connectome, with white matter signals reflecting information transmission between gray matter regions during seizures and structure-function coupling increasing during ictal periods.^34,35^ Our findings suggest that among patients with glioblastoma, those who develop TRE retain relatively greater white matter integrity, providing the structural substrate through which peritumoral hyperexcitability could propagate and synchronize into clinically manifest seizures. While non-tumor epilepsy has been characterized by paradoxical connectivity increases within specific circuits,^12–14^ the increased local efficiency across the majority of functional networks in TRE patients suggests that seizure susceptibility in glioblastoma may be more distributed, reflecting connectivity that is relatively broadly intact rather than regionally reorganized. This interpretation is consistent with the observation that network topology, specifically local efficiency, distinguished those with TRE more than raw connection strength, as locally well-connected neighborhoods may facilitate the synchronized neural recruitment required for seizure generation and spread.^34,35^ The absence of significant connectivity differences between TRE patients with and without focal-to-bilateral tonic-clonic seizures, or between those with drug-resistant and drug-responsive epilepsy, suggests that the connectivity profile associated with TRE was not appreciably driven by seizure severity or refractoriness, respectively, in this cohort. Similarly, connectivity associations were strongest when preoperative and postoperative seizures were combined into a single TRE phenotype rather than analyzed separately, suggesting that the relevant signal reflects a shared susceptibility to seizures rather than timing-specific mechanisms, though reduced statistical power in the subgroups cannot be excluded.

Contralateral global efficiency showed the strongest association with OS, and the consistent directional trend across all networks and hemispheres suggests the survival-relevant signal reflects overall brain reserve rather than a focal network effect. This is consistent with prior work showing that glioblastoma causes widespread connectome disruption extending beyond the ipsilesional hemisphere.^9,10^ The contralateral predominance suggests these metrics may capture the distant effects of tumor biology, given that glioblastoma alters white matter microstructure at greater distances from the tumor.^36^ Ipsilateral connectivity may be estimated less reliably near the lesion due to mass effect, making those associations more vulnerable to artifact. Our key survival findings, however, were contralateral, supporting the validity of these associations. Notably, no connectivity metric was associated with PFS, contrasting with prior work,^9^ suggesting that preserved white matter integrity may be more relevant to long-term survival than to the timing of initial tumor progression. This may reflect both the greater measurement variability inherent in radiographic progression assessment and a biological distinction between the determinants of initial treatment response and long-term survival.

Contralateral mean local efficiency was carried into mediation because it was significantly associated with both TRE and OS, the latter at nominal significance. Contralateral global efficiency, despite its significant association with OS, was not associated with TRE and therefore could not serve as a mediator of the seizure-survival relationship. Mediation analysis then revealed an asymmetry between the two causal directions tested. TRE did not mediate the connectivity-OS association, arguing against a framework in which seizures themselves confer the survival advantage. In the reverse direction, contralateral mean local efficiency partially mediated the TRE-OS association, though this depended on whether postoperative KPS was included in the outcome model. Because KPS could reflect downstream consequences of network integrity rather than independently confounding it, the model excluding KPS may better approximate the causal effect, though neither specification is definitive. Even under this specification, connectivity mediated only a small fraction of TRE’s survival advantage, indicating that preserved white matter could account for part but not most of the seizure-survival association. The remainder may operate through pathways not captured by structural networks, such as tumor biology, immune microenvironment, or treatment response. Together, this suggests that the connectivity-survival association may reflect an upstream factor, such as less aggressive tumor biology, while TRE retains prognostic value only partly explained by connectivity.

White matter integrity may also reflect broader clinical and demographic factors. FA-weighted connectivity metrics were negatively correlated with age and tumor volume and positively correlated with extent of resection and postoperative KPS, with KPS showing among the strongest associations (Supplementary Fig. 1). These correlations are intuitive, as younger patients have less age-related white matter degeneration, smaller tumors disrupt fewer tracts, and more circumscribed tumors permit more complete resections. These same clinical factors were also associated with both TRE and OS in glioblastoma, and we previously showed that smaller tumor volume, in particular, is associated with TRE.^8^ The attenuation of connectivity associations after covariate adjustment could therefore reflect this shared variance rather than a lack of biological relevance, arising from either true confounding or overadjustment, the latter consistent with the KPS sensitivity observed in our mediation analysis. The unadjusted and fully adjusted models may therefore represent upper and lower bounds of the true effect.

Several methodological choices warrant discussion. We retained perilesional ROIs within the graph rather than excluding them. Our lesion-aware tractography pipeline omits tumor and edema from atlas parcels and removes all streamlines passing through the tumor/edema mask, ensuring that no connectivity endpoints or pathways traverse the affected tissue. Consequently, perilesional nodes contribute their connectivity information, preserving the biologically relevant signal of tumor-related network disruption rather than artificially fragmenting the graph through node removal. This approach aligns with the conservative streamline exclusion strategy as previously described,^10^ while avoiding the parcellation failures inherent to surface-based methods in glioma patients through the use of template-based cortical atlases and deep learning-based subcortical segmentation. Moreover, survival and TRE models were adjusted for combined tumor and edema volume rather than enhancing tumor volume alone, which more accurately reflects the extent of connectome disruption.

This study has several limitations. The retrospective single-institution design limits generalizability, and the reliance on chart review for TRE classification may introduce misclassification, particularly for patients with unwitnessed or unreported seizures. The absence of a healthy control group means that all connectivity comparisons were relative within the glioblastoma cohort, as we could not determine whether the observed differences reflect preservation in TRE patients, disproportionate disruption in non-TRE patients, or both. The cross-sectional design, with connectivity measured at a single preoperative timepoint, precludes assessment of how network topology evolves with treatment, tumor progression, or seizure development. Our single-shell 30-direction diffusion acquisition at b=1000 s/mm^2^, while sufficient for SS3T-CSD tractography, provides lower angular resolution than multi-shell protocols, potentially reducing sensitivity to crossing fibers. Consequently, FA values in perilesional regions may be influenced by vasogenic edema, partial volume effects, or tumor infiltration even after streamline exclusion, and mass effect may alter FA through mechanical deformation rather than white matter pathology. In the clinical data, missing *MGMT* methylation status and KPS data reduced the sample available for adjusted survival analyses. Patients missing KPS data were older with shorter OS, and their exclusion may bias the adjusted cohort toward patients with better prognoses. Extent of resection was categorized from operative reports rather than quantified volumetrically from postoperative imaging, which could introduce imprecision, though the categorical measure was still significantly associated with OS in this cohort. Finally, we could not quantify the interval between symptom onset and diagnosis, so lead-time bias cannot be excluded as a contributor to the residual association between TRE and survival.

Structural connectivity metrics could serve as imaging biomarkers for both seizure risk stratification and survival prognostication in glioblastoma. Prospective validation would determine whether preoperative connectome analysis can complement existing prognostic models based on clinical and molecular features, and longitudinal imaging could establish whether connectivity changes over the disease course carry additional prognostic information. Integration with multimodal imaging and intraoperative electrophysiology could further clarify the relationship between peritumoral microstructure, network topology, and clinical outcomes, refining the structural account of the seizure-survival paradox these findings support.

## Acknowledgments

The authors are grateful to the patients and families who participated in this research. Computational analyses utilized Claude Opus (Anthropic) for assistance with Python code debugging and optimization. Funding for this work was provided by the National Institute of Neurological Disorders and Stroke (5T32NS091008-07 and 5T32NS091006-10 to D.Z., K99NS138680 to N.S.) and the American Epilepsy Society Research and Training Fellowship for Clinicians (1282834 to D.Z.).

## Author Contributions

DZ: study concept/design; methodology; software; data collection; analysis/interpretation of data; drafting/revising manuscript for content. MJ: methodology; software; analysis/interpretation of data; revising manuscript for content. JG (Gal): major role in acquisition of data. YF: major role in acquisition of data. AL: revising manuscript for content. QC: analysis/interpretation of data. CD: methodology; analysis/interpretation of data; major role in acquisition of data. MN: major role in acquisition of data; analysis/interpretation of data. JG (Garcia): major role in acquisition of data. RX: major role in acquisition of data. RP: revising manuscript for content. CE: major role in acquisition of data; analysis/interpretation of data; revising manuscript for content. SD: revising manuscript for content. JG (Gugger): revising manuscript for content. NS: revising manuscript for content. JS: revising manuscript for content. KD: methodology; resources; supervision; revising manuscript for content.

## Data Availability

De-identified data used in the present study are available from the corresponding author upon reasonable request, subject to institutional data sharing policies and collaborative approvals.

