## supplementary for "Structural Brain Networks and the Seizure-Survival Paradox in Glioblastoma"

### Supplementary Figures

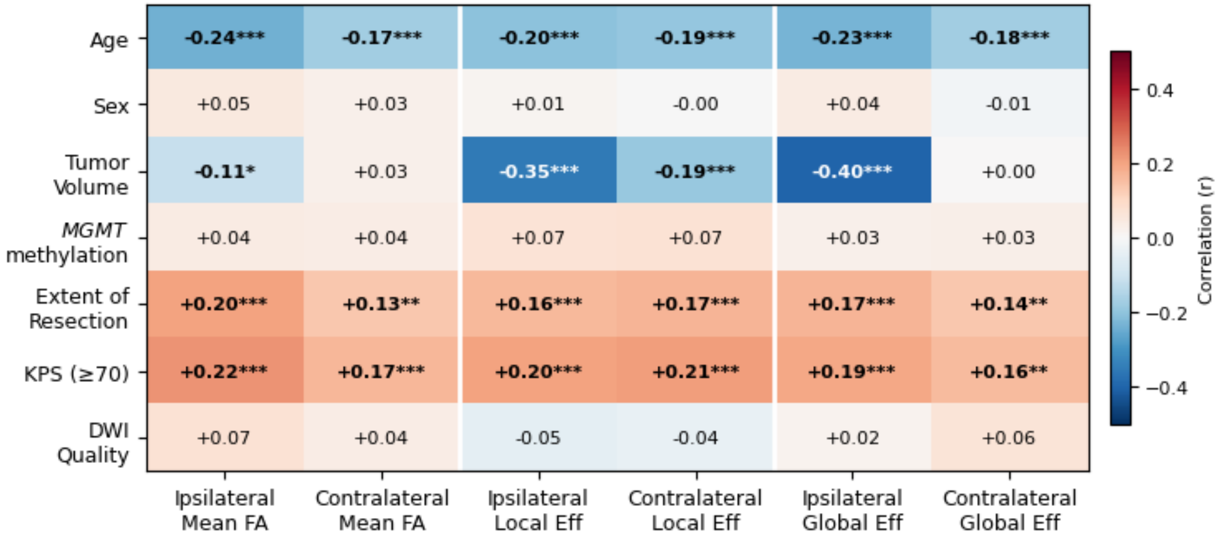


**Supplementary Figure 1.** Correlations between clinical covariates and fractional anisotropy (FA)-weighted connectivity metrics. Heatmap displays Spearman correlation coefficients between covariates (age, sex, tumor volume, *MGMT* methylation status, extent of resection, postoperative KPS, and DWI quality) and hemisphere-level connectivity metrics (mean FA, local efficiency, and global efficiency for ipsilateral and contralateral hemispheres). **p*<0.05, ***p*<0.01, ****p*<0.001.

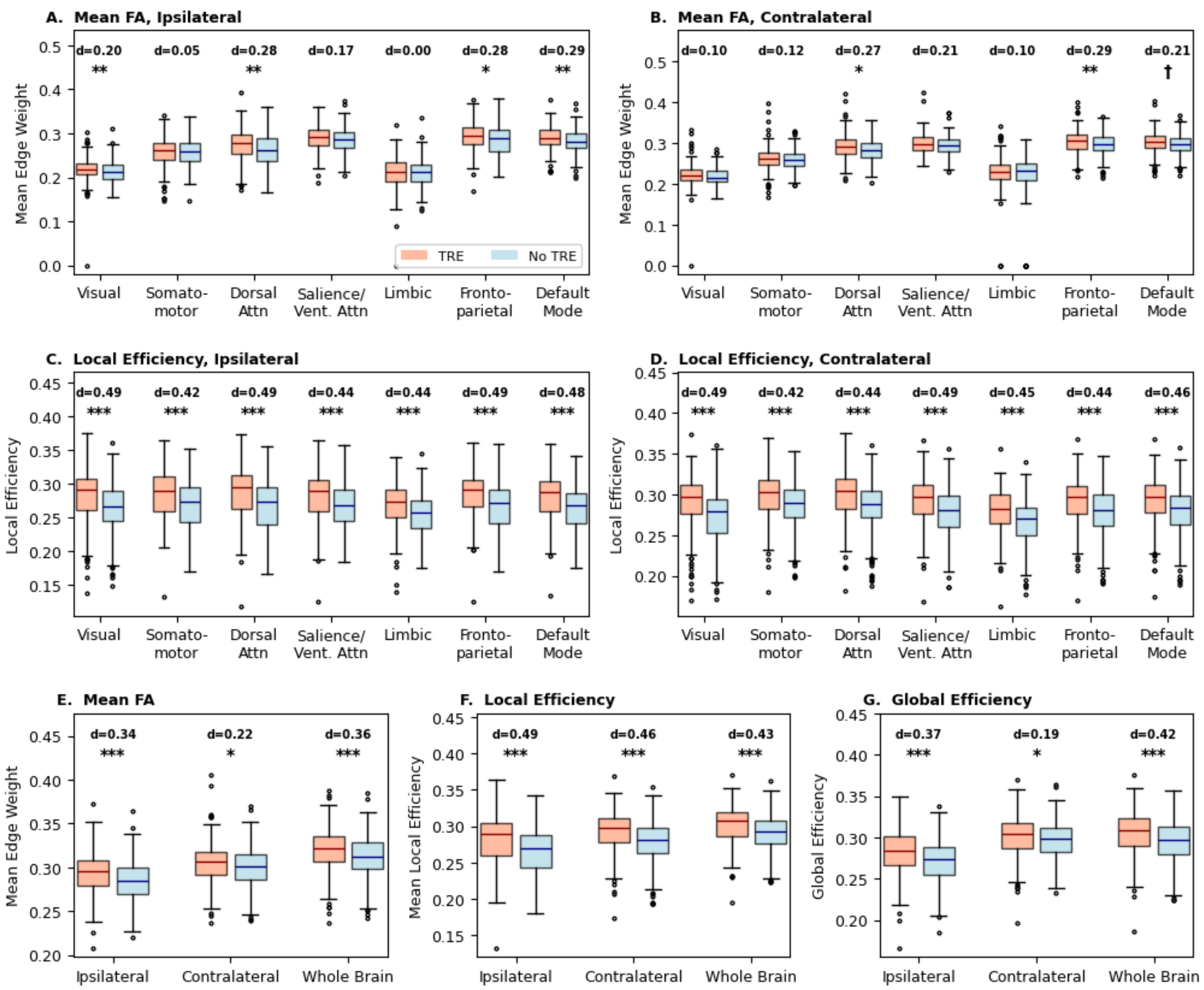


**Supplementary Figure 2.** Unadjusted fractional anisotropy (FA)-weighted connectivity metrics in patients with and without tumor-related epilepsy (TRE). (A-B) Mean FA edge weight per network for ipsilateral and contralateral hemispheres. (C-D) Local efficiency per network for ipsilateral and contralateral hemispheres. (E-G) Hemisphere-level summary metrics: mean FA (E), mean local efficiency (F), and global efficiency (G). Cohen's d effect sizes are shown above each comparison. Compared to adjusted models (Figure 2), unadjusted associations were stronger and more widespread. $*q<0.05$, $**q<0.01$, $***q<0.001$ (FDR-corrected), $^{\dagger}p<0.05$ (nominal).

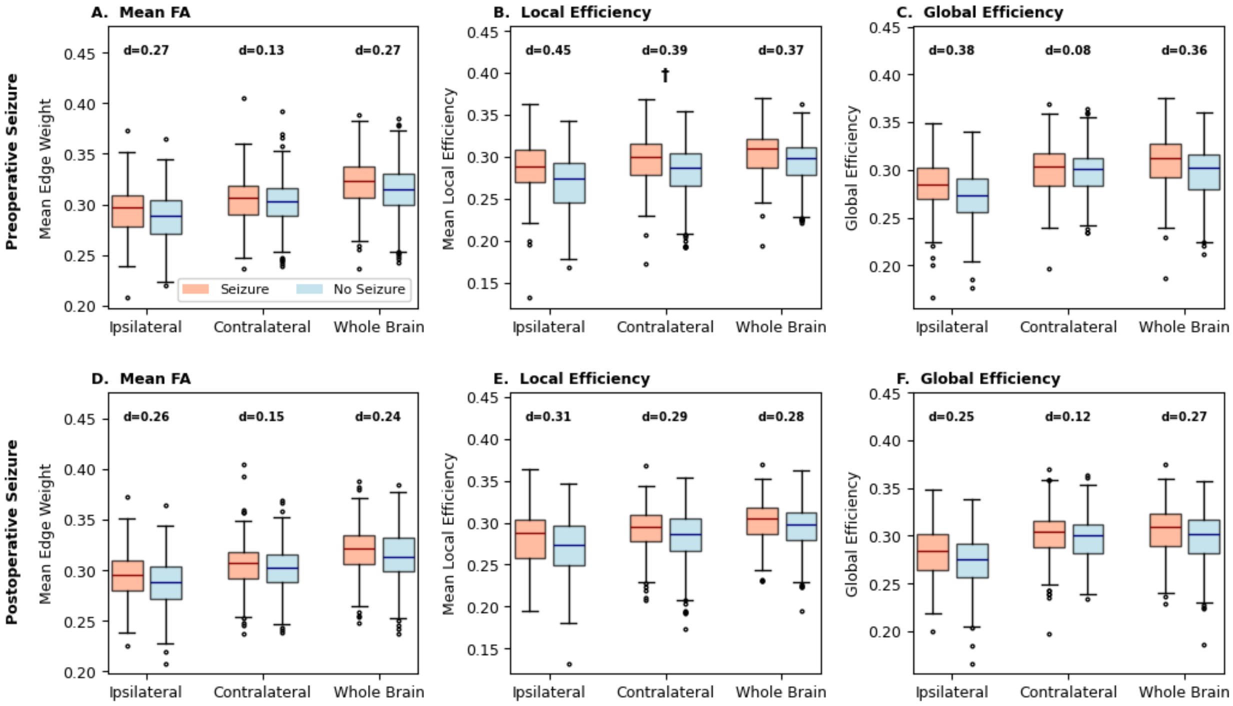


**Supplementary Figure 3.** Fractional anisotropy (FA)-weighted hemisphere-level connectivity metrics in preoperative and postoperative seizure subanalyses. Box plots compare distributions between patients with (red) and without (blue) seizures for ipsilateral, contralateral, and whole-brain regions. (A-C) Preoperative seizure subanalysis: mean FA edge weight (A), mean local efficiency (B), and global efficiency (C). (D-F) Postoperative seizure subanalysis (acute perioperative seizures excluded): mean FA edge weight (D), mean local efficiency (E), and global efficiency (F). Cohen's d effect sizes are shown above each comparison. Significance markers from logistic regression adjusting for age, sex, tumor volume, tumor location, and DWI quality (postoperative models additionally adjusted for extent of resection): *$q<0.05$, **$q<0.01$, ***$q<0.001$ (FDR-corrected), †$p<0.05$ (nominal).

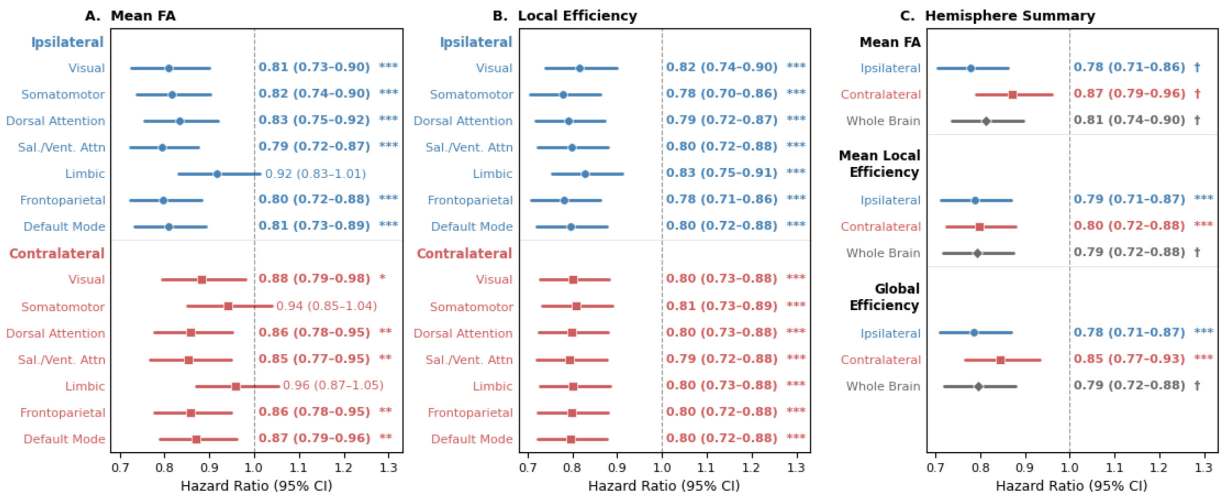


**Supplementary Figure 4.** Unadjusted associations between fractional anisotropy (FA)-weighted connectivity metrics and overall survival. Forest plots show hazard ratios (95% CI) from Cox proportional hazards regression without covariate adjustment. (A) Per-network mean FA edge weight. (B) Per-network local efficiency. (C) Hemisphere-level summary metrics including mean FA, mean local efficiency, and global efficiency for ipsilateral (blue), contralateral (red), and whole-brain regions (black). Hazard ratios are per 1 SD increase in connectivity metric. Compared to adjusted models (Figure 3), unadjusted associations were substantially stronger and more widespread. *$q<0.05$, **$q<0.01$, ***$q<0.001$ (FDR-corrected), $^{\dagger}p<0.05$ (nominal).

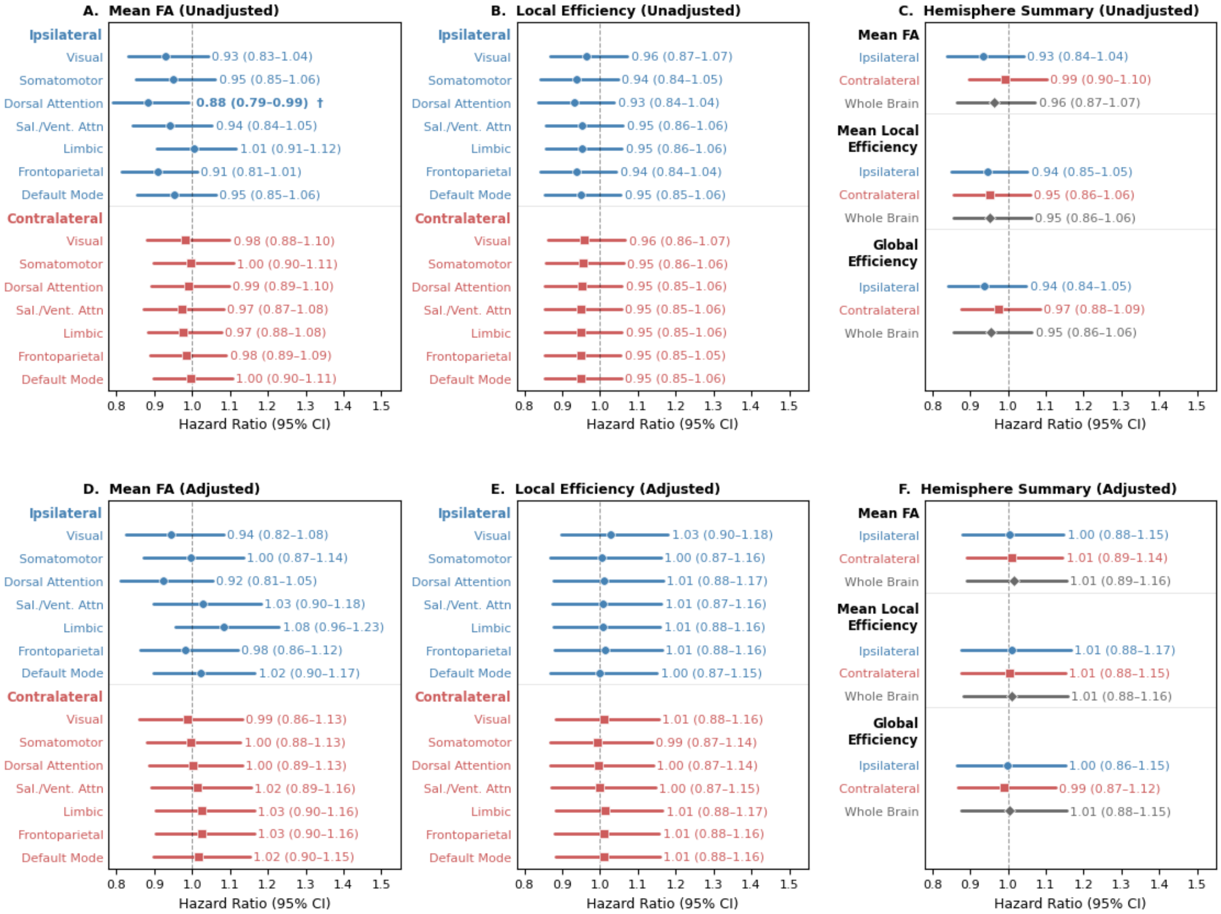


**Supplementary Figure 5.** Associations between fractional anisotropy (FA)-weighted connectivity metrics and progression-free survival. (A-C) Unadjusted models. (D-F) Adjusted models (age, sex, tumor volume, extent of resection, *MGMT* methylation status, postoperative KPS, and DWI quality). (A, D) Per-network mean FA edge weight in ipsilateral (blue) and contralateral (red) hemispheres. (B, E) Per-network local efficiency. (C, F) Hemisphere-level summary metrics. Hazard ratios are per 1 SD increase in connectivity metric. No significant associations were observed after FDR correction in either model. $^{\dagger}p<0.05$ (nominal).
